# Risk of Ischemic Heart Disease in Women with Dysmenorrhea: A Longitudinal Analysis of 251,264 Patients Across Three Diverse Cohorts

**DOI:** 10.1101/2024.10.04.24314831

**Authors:** Eugenia Alleva, Susan Khalil, Kimberly Glazer, Joanne Stone, Paola Viganò, Edgardo Somigliana, Stefan Konigorski, Isotta Landi, Chen Shengja, Ruchika Verma, Jannes Jagminat, Matteo Danieletto, Robert Hirten, Erwin Böttinger, Ipek Ensari, Thomas J. Fuchs, Leslee J. Shaw

## Abstract

This study explores the role of dysmenorrhea as a sex-specific ischemic heart disease (IHD) risk enhancing factor across three large cohorts of 251,264 individuals, two retrospective electronic health records cohorts, i.e., the Mount Sinai Health System, All of Us, and one prospective cohort, the Australian Longitudinal Survey on Women’s Health. Considering traditional and nontraditional young female-specific cardiovascular risk factors, hazard ratios for IHD were estimated through Cox regression models and propensity score matching, and dysmenorrhea was found to be significantly associated, with a 40% to 225% increase in the hazard of developing IHD. The risk was found to be higher in women of color and those with persistent dysmenorrhea beginning in adolescence. The addition of dysmenorrhea as a predictor beyond traditional cardiovascular risk scores improved risk stratification (AUROC 0.786 vs 0.798, p-value 0.02). Within the Mount Sinai’s electronic health records, we also found dysmenorrhea diagnostic codes to have a 37.3% false negative rate. To overcome the under-coding of the diagnosis, we implemented a large language model EHR-phenotyping algorithm which led to higher hazard ratios, together with self-reporting within the ALSWH (HR 2.5 and 3.3, respectively), compared to diagnostic code-only cohort definition (HR 1.5). These findings highlight the need to recognize dysmenorrhea’s impact beyond reproduction and call for greater clinical and research awareness.

## Introduction

Ischemic heart disease (IHD) is one of the main drivers of disease burden^1^ and a leading cause of death worldwide^2^. In the past decade, substantial declines in age-standardized disability-adjusted life-year and mortality rates in IHD have been observed globally. However, such improving trends have been more moderate for women compared to men^1,3^

In the cardiovascular health space, sex- and gender-based inequalities exist and disproportionally affect women who identify with minoritized races and ethnicities^4^. Screening and diagnostic pathways report lower accuracy at guiding preventive care in women compared to men^5^ and traditional risk assessment approaches show sub-optimal performance in women because largely derived from male populations^6^. Therefore, the early identification of at-risk individuals and an effective primary prevention are a priority for women, and sex-specific risk markers are needed. To this aim, adverse pregnancy outcomes, such as gestational hypertension, have recently been included in the list of cardiovascular risk enhancing features in the American College of Cardiology/American Heart Association guidelines for primary cardiovascular disease prevention^7^. Nevertheless, given our limited knowledge of the cardiovascular sex-specific risk factors from a woman’s reproductive years^5,7,8^, the promotion of cardiovascular health in women cannot be limited to pregnancy alone.

Menstrual-related conditions, such as irregular menstrual cycles, polycystic ovary syndrome, and heavy menstrual bleeding, have emerged as potential enhancers of IHD risk^9–11^. Among them, dysmenorrhea, i.e., the cyclical menstruation-related pain, holds unprecedent promises in that it is associated with known contributors to atherosclerotic cardiovascular disease (ASCVD), such as release of inflammatory mediators^12^, autonomic nervous system alterations^13,14^, and stress^15,16^. Although affecting up to 30% of menstruating individuals in its severe form^12^, dysmenorrhea is still a greatly underexplored risk indicator, because it has only been associated to cardiovascular outcomes, i.e., IHD^17^ and stroke^18^, as a stand- alone factor.

Leveraging three longitudinal cohorts diverse in terms of self-reported race and ethnicity, we demonstrated the role of dysmenorrhea as a sex-specific IHD risk enhancing factor. To this end, we considered other menstrual-related conditions and traditional cardiovascular risk factors as covariates and examined how this association varied across different dysmenorrhea case definition methods. Moreover, we showcased the valence of dysmenorrhea in the assessment of IHD risk, we tested the prognostic value of dysmenorrhea beyond traditional cardiovascular risk models.

We acknowledge that the investigation of sex-specific reproductive health indicators is only the tip of the iceberg for the improvement of women’s cardiovascular health, because gender and social aspects also need to be considered for the equitable stratification of IHD risk. Nevertheless, this work aims at contributing to the list of risk factors typical of a woman’s reproductive lifespan beyond pregnancy, thus improving the cardiovascular health of vulnerable populations at highest need for better risk stratification and targeted primary prevention.

## Results

### Cohorts Description

In this study we used three diverse longitudinal datasets totaling 251,264 distinct patients. Two large electronic health record (EHR) datasets, the All of Us Research Program [AoU] and Mount Sinai [MS] EHR, and a prospective longitudinal study, the Australian Longitudinal Survey on Women’s Health [ALSWH] (See Figure 1 Panel A). Dysmenorrhea cases were defined using three distinct approaches, building a total of five distinct cohorts (Figure 1, Panel A): (1) SNOMED-coded diagnoses alone (Aou and MS Full); (2) a large language model (LLM) EHR phenotyping algorithm (Figure 1 Panel B) that leverages both coded diagnoses and clinical notes (MS LLM-EHR); and (3) self-reported dysmenorrhea from the ALSWH. A separate ALSWH-S5 sub-cohort was built from the ALSWH dataset using survey 5 (S5) as the index date, where participant age at S5 is comparable to the MS Full, MS LLM-EHR and AoU cohorts (See Table 1).

**Figure 1:**
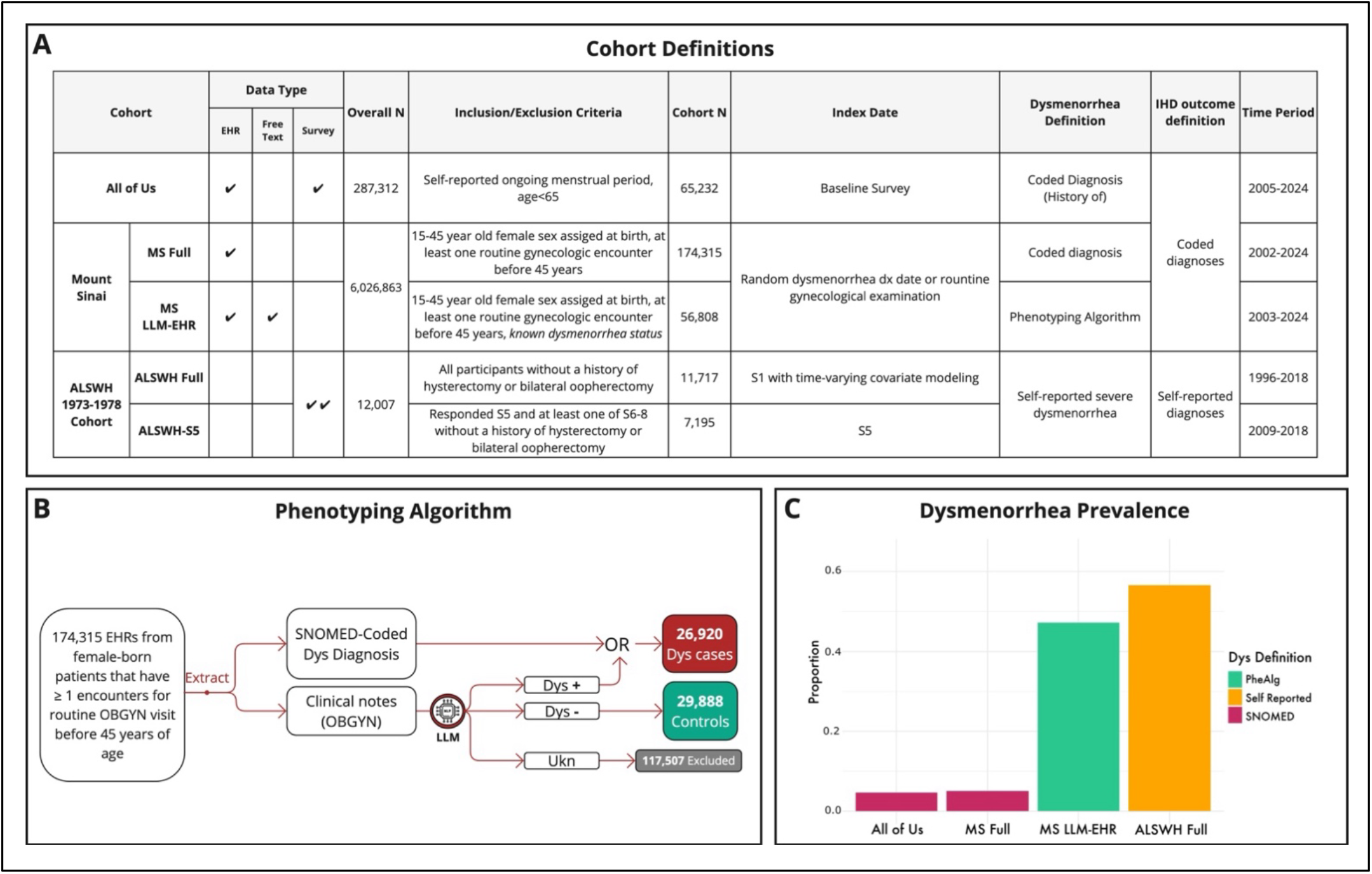
Cohorts Overview. Panel A: Cohort Definition Methods. We used three datasets, All of Us, Mount Sinai EHR and ALSWH to build five cohorts with varying dysmenorrhea definition methods. Panel B: Phenotyping Algorithm Overview. A combination of coded diagnoses and LLM-based clinical note classification was used to identify ascertained dysmenorrhea cases and controls. Patients with neither a coded diagnosis, nor at least one clinical note classified by the LLM as dysmenorrhea present or absent was defined considered unknown dysmenorrhea status (Unk) and excluded from the MS LLM-EHR cohort. Panel C: Prevalence of Dysmenorrhea Across Cohorts. Prevalence of dysmenorrhea using coded diagnoses alone is considerably lower than what is estimated using self-report or phenotyping algorithm. Prevalence for the ALSWH is ascertained as having ever reported dysmenorrhea across S1-S8 surveys. Abbreviations - Dys: dysmenorrhea; Unk: Unknown dysmenorrhea status, LLM: language model; PheAlg: Phenotyping Algorithm.

**Table 1:** Covariate Distribution Across Cohorts. Mean (SD) are reported for continuous variables, percentages (N) are reported for categorical variables. AoU: All of Us. MS Full: Mount Sinai cohort using only SNOMED codes for dysmenorrhea case definition. MS LLM-EHR: Mount Sinai cohort using phenotyping algorithm for dysmenorrhea case definition and excluding patients with unknown dysmenorrhea status. ALSWH Full: Australian Longitudinal Survey on Women’s Health cohort including data from all surveys with covariate updated at each survey with new responses. BMI is reported as average BMI across surveys for each participant. Categorical covariates are reported as presence at any survey for each participant. ALSWH S5: ALSWH Survey 5 cohort. Covariate values is determined from survey 5 alone. Dys: Dysmenorrhea.

| Cohort | Mount Sinai LLM-EHR |  | Mount Sinai Full |  | All of Us |  | ALSWH Full |  | ALSWH S5 |  |
| --- | --- | --- | --- | --- | --- | --- | --- | --- | --- | --- |
|  | Dys, N = | Control, N = | Dys, N = | Control, N = | Dys, N = | Control, N = | Dys, N = | Control, N = | Dys, N = | Control, N = |
| **Variable** | 26,920 | 29,888 | 8,788 | 165,527 | 3,069 | 62,163 | 6,562 | 5,155 | 1,506 | 5,689 |
| **Age** | 31.40<br>(7.47) | 32.63<br>(6.66) | 30.27<br>(7.51) | 32.57<br>(6.83) | 34.89<br>(14.89) | 35.34 (14.80) | 20.28<br>(1.50) | 20.35<br>(1.49) | 33.27<br>(1.49) | 33.21<br>(1.47) |
| **Race** |  |  |  |  |  |  |  |  |  |  |
| Asian | 6.0%<br>(1,622) | 9.3%<br>(2,772) | 5.9%<br>(521) | 8.0%<br>(13,288) | 2.4% (73) | 4.6% (2,829) | N/A | N/A | N/A | N/A |
| Black | 10.0%<br>(2,691) | 7.5%<br>(2,256) | 13%<br>(1,164) | 8.4%<br>(13,967) | 20% (611) | 19% (11,527) | N/A | N/A | N/A | N/A |
| Hispanic | 13%<br>(3,488) | 10%<br>(2,995) | 15%<br>(1,328) | 10%<br>(17,119) | 23% (706) | 25% (15,766) | N/A | N/A | N/A | N/A |
| Mixed/Other | 6.9%<br>(1,854) | 6.9%<br>(2,064) | 7.3%<br>(642) | 7.3%<br>(12,148) | 7.3% (225) | 8.1% (5,042) | N/A | N/A | N/A | N/A |
| Unknown | 20%<br>(5,456) | 27%<br>(7,947) | 20%<br>(1,765) | 26%<br>(43,390) | 0.9% (28) | 1.1% (669) | N/A | N/A | N/A | N/A |
| White | 44%<br>(11,809) | 40%<br>(11,854) | 38%<br>(3,368) | 40%<br>(65,615) | 46%<br>(1,426) | 42% (26,330) | N/A | N/A | N/A | N/A |
| **Irregular Menstruation** | 8.3%<br>(2,238) | 5.1%<br>(1,530) | 9.1%<br>(802) | 4.9%<br>(8,121) | 38%<br>(1,178) | 9.0% (5,614) | 59%<br>(3,874) | 36%<br>(1,854) | 37%<br>(552) | 13%<br>(759) |
| **Heavy Menstrual Bleeding** | 6.2%<br>(1,674) | 2.6%<br>(781) | 8.5%<br>(744) | 2.5%<br>(4,206) | 22% (663) | 3.5% (2,193) | 71%<br>(4,642) | 24%<br>(1,231) | 63%<br>(949) | 9.6%<br>(546) |
| **Endometriosis** | 2.4%<br>(635) | 0.5%<br>(137) | 3.5%<br>(307) | 0.8%<br>(1,275) | 14% (418) | 2.4% (1,481) | 12%<br>(767) | 2.9%<br>(152) | 15%<br>(225) | 4.8%<br>(271) |
| **Fibroids** | 4.4%<br>(1,178) | 2.1%<br>(618) | 5.1%<br>(451) | 2.6%<br>(4,282) | 17% (518) | 5.6% (3,464) | N/A | N/A |  |  |
| **PCOS** | 2.8%<br>(745) | 2.1%<br>(625) | 2.8%<br>(247) | 2.2%<br>(3,624) | 5.8% (177) | 2.2% (1,346) | N/A | N/A | 9.1%<br>(137) | 5.0%<br>(285) |
| **Migraine** | 9.4%<br>(2,533) | 5.8%<br>(1,741) | 11%<br>(999) | 6.2%<br>(10,296) | 33% (999) | 11% (6,935) | 86%<br>(5,618) | 72%<br>(3,709) | 88%<br>(1,328) | 78%<br>(4,421) |
| **Hormonal Contraception** | 17%<br>(4,696) | 21%<br>(6,259) | 14%<br>(1,267) | 17%<br>(28,038) | 53%<br>(1,633) | 20% (12,458) | 88%<br>(5,749) | 88%<br>(4,549) | 24%<br>(365) | 36%<br>(2,046) |
| **Ever Smoker** | 12%<br>(3,119) | 9.4%<br>(2,817) | 11%<br>(985) | 9.5%<br>(15,659) | 12% (370) | 13% (8,271) | 56%<br>(3,677) | 48%<br>(2,492) | 44%<br>(658) | 38%<br>(2,157) |
| **Hypertension** | 2.2%<br>(600) | 1.7%<br>(509) | 2.1%<br>(184) | 1.9%<br>(3,222) | 24% (744) | 15% (9,256) | 12%<br>(819) | 9.0%<br>(464) | 11%<br>(162) | 7.9%<br>(448) |
| **Hypertensive Disorders of Pregnancy** | 0.6%<br>(151) | 0.6%<br>(182) | 0.4%<br>(35) | 0.7%<br>(1,206) | 6.3% (192) | 3.5% (2,153) | N/A | N/A | 8.6%<br>(129) | 6.3%<br>(357) |
| **Diabetes** | 1.4%<br>(381) | 1.3%<br>(391) | 1.6%<br>(137) | 1.4%<br>(2,277) | 14% (435) | 9.3% (5,790) | 3.5%<br>(230) | 2.3%<br>(118) | 2.0%<br>(30) | 1.4%<br>(78) |
| **Gestational Diabetes** | 0.6%<br>(151) | 0.7%<br>(205) | 0.3%<br>(28) | 0.9%<br>(1,482) | 2.8% (87) | 2.2% (1,387) | N/A | N/A | 3.1%<br>(47) | 2.4%<br>(137) |
| **BMI** | 25.33<br>(5.91) | 24.74<br>(5.52) | 25.29<br>(6.04) | 24.99<br>(5.69) | 30.86<br>(11.00) | 30.06 (10.34) | 24.85<br>(5.12) | 24.15<br>(4.62) | 26.60<br>(6.36) | 25.67<br>(5.67) |
| **IHD** | 0.3%<br>(75) | 0.1%<br>(36) | 0.3%<br>(24) | 0.2%<br>(281) | 1.8% (54) | 0.7% (439) | 1.3%<br>(83) | 1.0% (54) | 1.5%<br>(22) | 0.7%<br>(40) |
| **Years of Follow-Up** | 4.20<br>(3.38) | 3.80<br>(3.30) | 4.06<br>(3.47) | 3.60<br>(3.20) | 2.72 (1.32) | 2.77 (1.27) | 18.53<br>(5.62) | 17.05<br>(6.44) | 8.11<br>(1.85) | 8.13<br>(1.84) |

Descriptive statistics and distribution of covariates across the five cohorts are presented in Table 1. A total of 1042 IHD events occurred across cohorts: MS Full (305 events, incidence rate of 0.0005/person- year), MS LLM-EHR (111 events, incidence rate of 0.0005/person-year), AoU (493 events, incidence rate of 0.0027/person-year), ALSWH Full (137 events, incidence rate of 0.0007/person-year) and ALSWH-S5 (62 events, 0.0011/ person-year). As anticipated, patients with dysmenorrhea exhibited a higher prevalence of endometriosis, fibroids, irregular menstruation (IM), and heavy menstrual bleeding (HMB) across all cohorts.

Prevalence of dysmenorrhea was 4.6% in the MS Full and 5.0% in the AoU cohorts, both based on coded diagnoses, compared to 56.6% from self-reporting in ALSWH. Using the phenotyping algorithm on the MS LLM-EHR cohort, the estimated prevalence in the Mount Sinai patient population rose to 47.2%, and the use of SNOMED codes alone in this cohort had a false negative rate of 37.3%. The dysmenorrhea LLM-EHR phenotyping algorithm had an overall performance of 0.88 F1 score (precision = 0.90, recall = 0.87), while the language model component alone of 0.90 F1 score (precision = 0.91, recall = 0.88) (Full results can be found in Appendix II).

Compared to the AoU cohort, the MS cohorts exhibited a lower prevalence of comorbidities, including diabetes and hypertension, and a significantly lower mean BMI. Importantly, covariate distribution did not significantly change between MS Full and MS LLM-EHR cohorts, suggesting that the phenotyping algorithm did not introduce a selection bias (See Appendix III).

### Primary Analyses: IHD Risk

Table 2 reports the hazard ratios (HR) estimated using both full propensity score matching with weighted Cox regression, as well as multivariate Cox regression models that incorporate an increasing number of covariates.

**Table 2:**
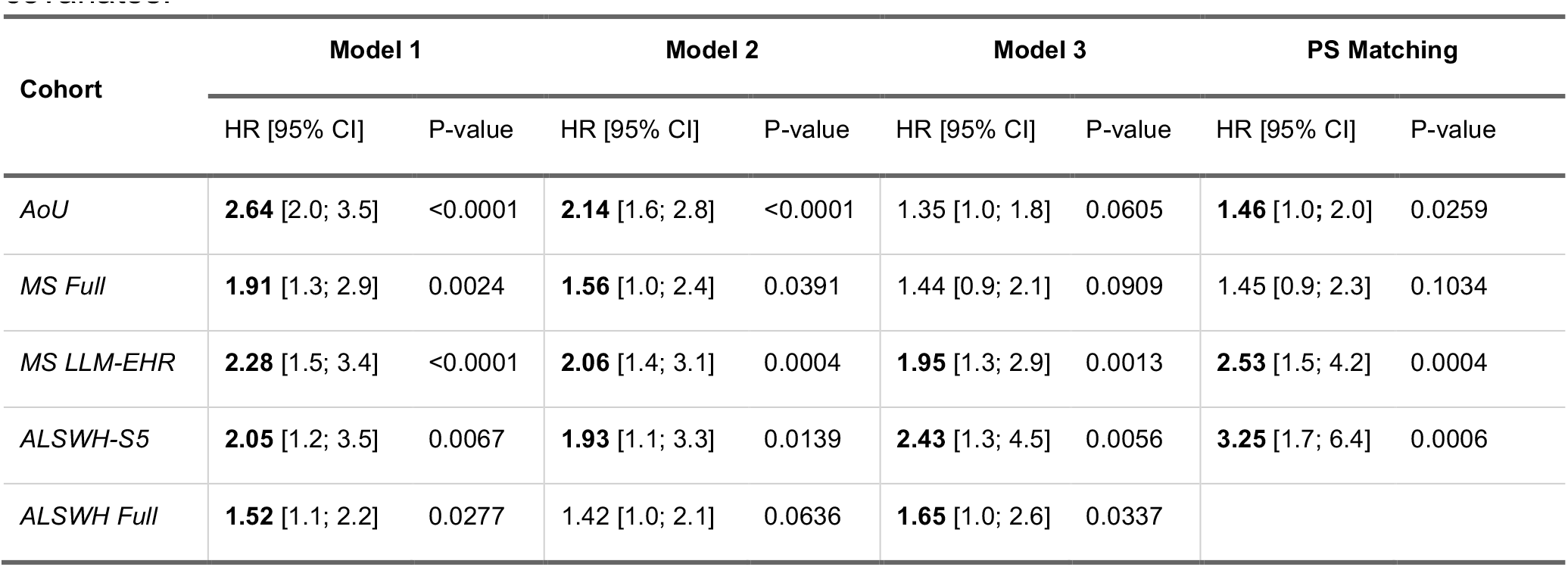
Hazard Ratio (HR) [95% Confidence Intervals] of Ischemic Heart Disease Across Cohorts. Model 1: Dysmenorrhea only. Model 2: Addition of BMI, Smoking, Hypertension, Diabetes and Race and ethnicity. Model 3: Addition of hormonal contraceptive use, endometriosis, fibroids, irregular menstruation, heavy menstrual bleeding, PCOS, migraine, gestational diabetes, hypertensive disorder of pregnancy.

| Cohort | Model 1 |  | Model 2 |  | Model 3 |  | PS Matching |  |
| --- | --- | --- | --- | --- | --- | --- | --- | --- |
|  | HR [95% CI] | P-value | HR [95% CI] | P-value | HR [95% CI] | P-value | HR [95% CI] | P-value |
| *AoU* | **2.64** [2.0; 3.5] | <0.0001 | **2.14** [1.6; 2.8] | <0.0001 | 1.35 [1.0; 1.8] | 0.0605 | **1.46** [1.0; 2.0] | 0.0259 |
| *MS Full* | **1.91** [1.3; 2.9] | 0.0024 | **1.56** [1.0; 2.4] | 0.0391 | 1.44 [0.9; 2.1] | 0.0909 | 1.45 [0.9; 2.3] | 0.1034 |
| *MS LLM-EHR* | **2.28** [1.5; 3.4] | <0.0001 | **2.06** [1.4; 3.1] | 0.0004 | **1.95** [1.3; 2.9] | 0.0013 | **2.53** [1.5; 4.2] | 0.0004 |
| *ALSWH-S5* | **2.05** [1.2; 3.5] | 0.0067 | **1.93** [1.1; 3.3] | 0.0139 | **2.43** [1.3; 4.5] | 0.0056 | **3.25** [1.7; 6.4] | 0.0006 |
| *ALSWH Full* | **1.52** [1.1; 2.2] | 0.0277 | 1.42 [1.0; 2.1] | 0.0636 | **1.65** [1.0; 2.6] | 0.0337 |  |  |

In the multivariate Cox regression models, we incrementally included covariates to adjust for potential confounding factors. Model 1 included dysmenorrhea only, Model 2 added cardiovascular risk factors BMI, smoking, hypertension, diabetes, and race and ethnicity, and Model 3 further incorporated all other covariates: hormonal contraceptive use, endometriosis, fibroids, irregular menstruation, heavy menstrual bleeding, PCOS, m^16^igraine, gestational diabetes and hypertensive disorders of pregnancy. The full results, including detailed estimates for all covariates are provided in Table C 3: Prevalence of IHD Appendix IV.

For all but the MS Full cohort, dysmenorrhea was found significantly associated with a 40% to 225% increase in the hazard of developing IHD when estimated via propensity score matching and weighted regression. Interestingly, HR estimated using SNOMED coded diagnoses alone were lower than those estimated using either LLM-EHR phenotyping algorithm or self-reported dysmenorrhea on the ALSWH and MS LLM-EHR cohorts (MS Full HR=1.40 [0.9-2.3], AoU HR=1.42 [1-2], versus MS LLM-EHR HR=2.53 [1.5-4.2], ALSWH-S5 HR=3.25 [1.7-6.4], see Table 2).

### Secondary Analyses: Effect Heterogeneity

#### Race and Ethnicity

To investigate whether dysmenorrhea is differentially associated with IHD across different racial and ethnic groups, we performed stratified analyses in the AoU, MS Full, and MS LLM-HER cohorts across aggregated racial and ethnic groups (White vs. Black, Indigenous and People of Color [BIPOC]) as well as by each racial and ethnic group separately (See Appendix V). We created a BIPOC category inclusive of all racial and ethnic groups excluding White self-reported race and including Asian and Hispanic self- reported race and ethnicity for sufficient statistical power in analyses.

Figure 2, Panel A presents the estimated HR for the aggregated racial and ethnic categories across each cohort. We observed an association between dysmenorrhea and increased IHD risk in BIPOC participants and no association among White patients with dysmenorrhea. This differential association across race and ethnicity was further supported by a statistically significant interaction term in adjusted regression models on the MS LLM-EHR (p-value 0.02) and AoU (p-value 0.04) cohorts, albeit this interaction was not significant in MS Full or when propensity score matching was used. It is important to note that the incidence of IHD was considerably lower among White participants compared to BIPOC participants (incidence of 2.3/10^4^py [MS LLM-EHR], 2.2/10^4^py [MS Full], and 18.1/10^4^py [AoU] in White participants vs. 10.3/10^4^py [MS LLM-EHR], 9.4/10^4^py [MS Full], and 33.3/10^4^py [AoU] in BIPOC participants), which contributes to the higher uncertainty in the HR estimates for the White group. We additionally tested the potential confounding effect of socio-economic variables (education, income and insurance type) on this interaction on the AoU dataset, demonstrating no significant confounding (See Appendix VI and Supplementary Material II).

**Figure 2:**
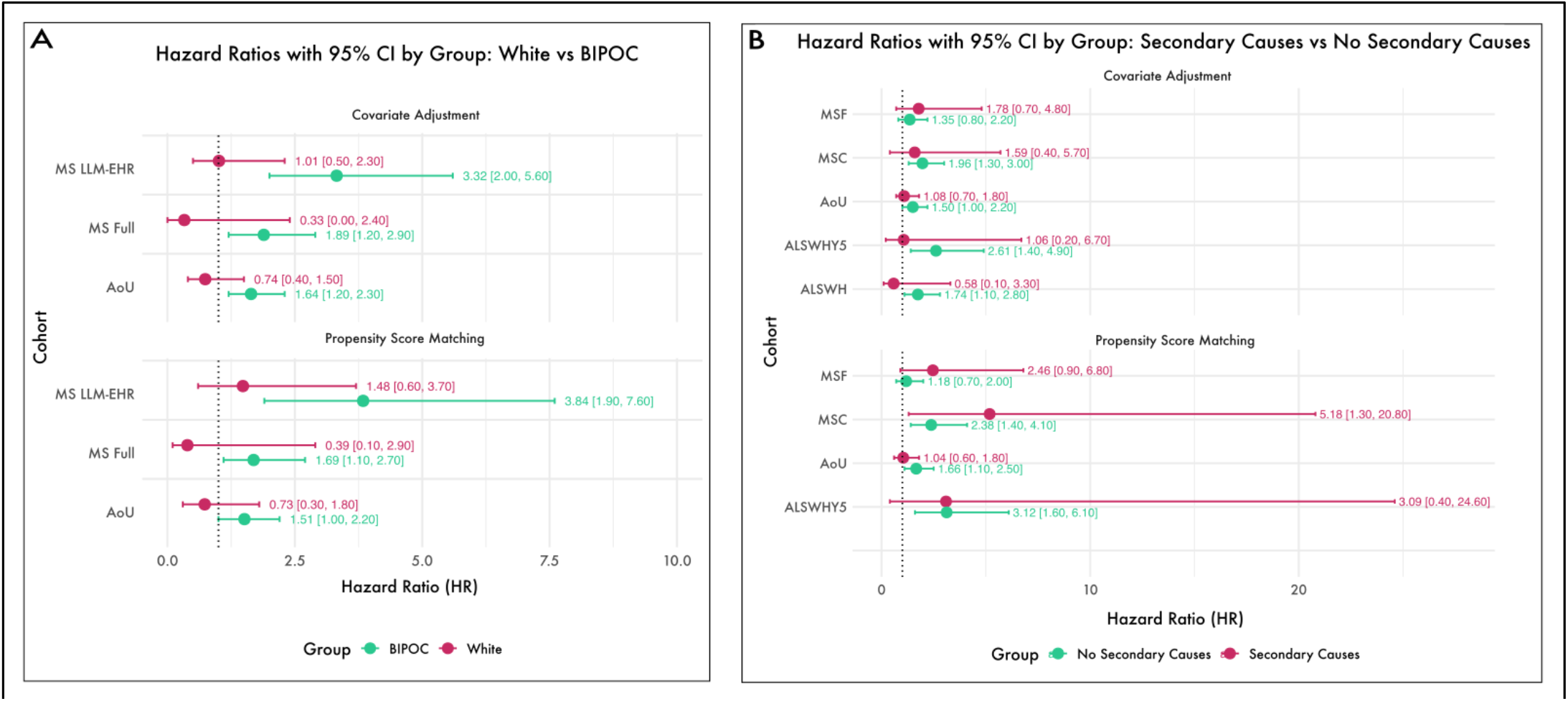
Secondary Analyses – Effect Heterogeneity. Panel A: Hazard ratios (HR) with 95% confidence intervals (CI) for IHD risk stratified by aggregated racial and ethnic groups (White vs. Black, Indigenous, and People of Color [BIPOC]). The analyses are presented for both covariate adjustment and propensity score matching across different cohorts (MS Full, MS LLM-EHR, and AoU). Panel B: Hazard ratios (HR) with 95% confidence intervals (CI) for IHD risk stratified by the presence or absence of secondary causes of dysmenorrhea, specifically fibroids and endometriosis. The analyses are similarly conducted using both covariate adjustment and propensity score matching methods across the same cohorts (MS Full, MS LLM-EHR, AoU, and ALSWH-S5). Green points indicate HR estimates for the White group (Panel A) or participants without secondary causes of dysmenorrhea (Panel B), while pink points represent the BIPOC group (Panel A) or participants with secondary causes (Panel B). BIPOC: Black, Indigenous and People of Color.

Full results, including racial and ethnic group-specific HR estimates can be found in Appendix V and Supplementary Material II. We observed a pattern of increased IHD risk for Hispanic (AoU HR=1.95 [1.2-3.3 p-value=0.01], MS LLM-EHR HR=3.14 [1.3-7.7, p-value=0.01], MS Full HR=1.8 [1.0-3.2, p-value=0.05]) and Asian patients with dysmenorrhea (MS Full HR=7.31 [1.4-37.4, p-value=0.02], MS LLM- EHR HR=18.77 [2.0-172.4, p-value=0.01]) across cohorts, but no association among White patients.

#### Primary vs Secondary Dysmenorrhea

To assess whether secondary causes of dysmenorrhea, such as endometriosis and fibroids, influence the association between dysmenorrhea and IHD, we conducted stratified analyses across all cohorts, comparing participants with and without secondary causes of dysmenorrhea. The results are detailed in Table 3, Panel B. Our findings indicate that the presence of secondary causes does not significantly modify the association between dysmenorrhea and IHD. The HR estimated within each stratum are consistent across cohorts, with no discernible trend suggesting a moderating effect. Furthermore, interaction models incorporating terms for the presence of secondary causes alongside dysmenorrhea did not yield significant interaction effects across any cohort or model specification. Comprehensive results, including interaction analyses, are provided in Appendix V and Supplementary Material II.

#### Age-dependent Effect

To investigate if the effect of dysmenorrhea varies with age, we estimated the time-varying coefficient for dysmenorrhea using age as a timescale for cox regression modeling in the ALSWH cohort, as depicted in Figure 3, Panel A. Additionally, we performed piecewise regression for the age groups ≤30, >30 to <40, and ≥40. Our results reveal an inverted U-shaped pattern, with a peak HR of 2.27 [1.2-4.2] (p-value = 0.008) in patients aged 30-40, compared to an HR of 1.06 [0.5-2.3] (p-value = 0.879) in those aged < 30, and an HR of 1.58 [0.7-3.4] (p-value = 0.240) in those aged ≥40. However, these findings are not confirmed in the MS LLM-EHR and MS Full cohorts, where participants aged 30-40 show similar HR with respect to participants <30 or =40 (HR = 2.58 [1.4-4.7] for age <30 or =40 vs. HR = 2.48 [1.0-6.3] for age 30-40 in MS LLM-EHR cohort and HR = 1.41 [0.8-2.5] for age <30 or =40 vs. HR = 1.55 [0.7-3.3] for age 30-40 in MS Full cohort).

**Figure 3:**
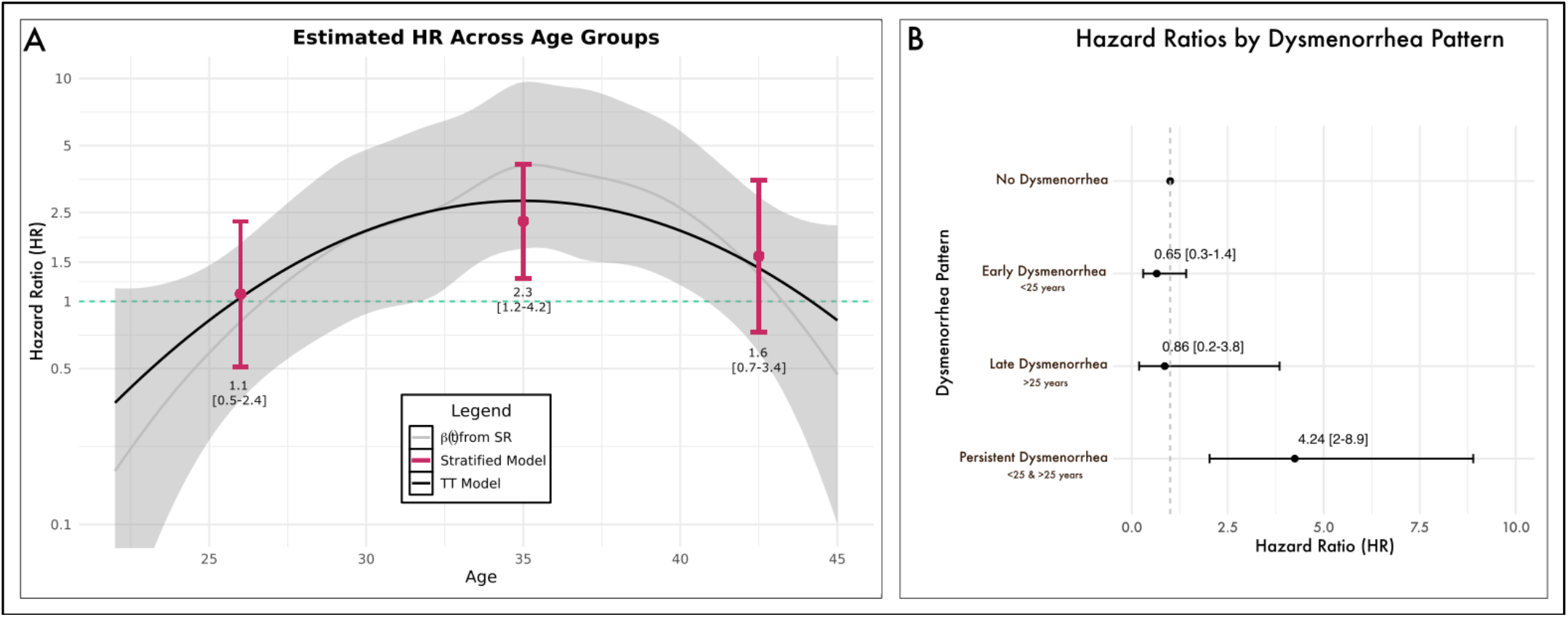
Age at Dysmenorrhea and IHD Risk. Panel A: Hazard ratios (HR) for IHD risk across different age groups at the onset of dysmenorrhea. The grey line represents the estimated time-varying coefficient derived from Schoenfeld Residuals (SR). The black line illustrates the polynomial time-transformed dysmenorrhea indicator, and the pink points with error bars indicate the HR estimates from a stepwise regression model stratified by age groups (<30, 30-40, >40 years). Panel B: Hazard ratios for IHD risk based on specific dysmenorrhea patterns. The analysis compares no dysmenorrhea, early dysmenorrhea onset (<25 years), late dysmenorrhea onset (≥25 years), and persistent dysmenorrhea (onset <25 years with continued symptoms into later age) using weighted cox regression after full propensity score matching. The HR estimates are presented with 95% confidence intervals (CI). SR = Schoenfeld Residuals; TT = Time-transformed. SR = Schoenfeld Residuals. TT = Time-transformed.

#### Natural History of Dysmenorrhea

To assess if patients with different natural history patterns of dysmenorrhea have varying IHD risks, we categorized patients of the ALSWH-S5 cohort into three groups: persistent dysmenorrhea (dysmenorrhea present both before age 25 and at the index date, N=937), early dysmenorrhea (present only before age 25, N=1855), and late dysmenorrhea (present at the index date but absent before age 25, N=569). We estimated the HR associated with each natural history pattern separately (Figure 3, Panel B). Women with persistent dysmenorrhea had a significantly increased IHD risk of more than four times that of women without dysmenorrhea (HR=4.24 [2.0-8.9], p-value <0.001 for weighted cox regression after full propensity score matching, HR= 2.96 [1.5-5.9], p-value 0.002 for multivariate cox regression [See Supplementary Material II for full results]), whereas women with early or late dysmenorrhea do not show a significant increase in IHD risk. Moreover, when compared with patients with early or late dysmenorrhea pattern, women with persistent dysmenorrhea have a HR of 4.26 [2.0-9.05] (p-value <0.001) (See Appendix V and Supplementary Material II for more results).

On the AoU cohort, we performed a cox regression limited to patients with dysmenorrhea and stratified for first diagnosis <25 vs >25 years of age. The results demonstrate a statistically significant HR of 1.86 [1.1-3.2] (p-value = 0.025) for first diagnosis before age 25 (N=1217), compared to an HR of 1.21 [0.8- 1.7] (p-value = 0.291) for first diagnosis after age 25 (N=1840). Moreover, when regressing scaled dysmenorrhea diagnosis age on IHD in this sub-cohort, we observed an inverse relationship between age at first diagnosis and IHD hazard, as indicated by the significantly decreased HR for continuous normalized age at first diagnosis (HR: 0.65 [0.5-0.9], coefficient = -0.430, p-value = 0.018).

### Prognostic Value of Dysmenorrhea

We compared a baseline IHD risk prediction model using the Framingham Offspring 30-year CVD risk score (30FRS) as a predictor to an expanded model additionally including dysmenorrhea as a predictor to assess prognostic value of dysmenorrhea. We compared baseline and extended models in terms of discriminative power (C-index), goodness-of-fit (via log-likelihood ratio test [LRT] for nested models), and continuous net reclassification index (cNRI) (Figure 4, Panel A). Separate stratified analyses across aggregated racial and ethnic groups were also performed. The MS LLM-EHR cohort was used for this analysis. Because the 30FRS was developed for predicting risk for a combination of cardiovascular disease (CVD) outcomes beyond IHD, including atrial fibrillation, stroke, peripheral vascular disease and heart failure, we additionally performed comparable analyses on combined CVD outcome predictions (Figure 4, Panels B).

**Figure 4:**
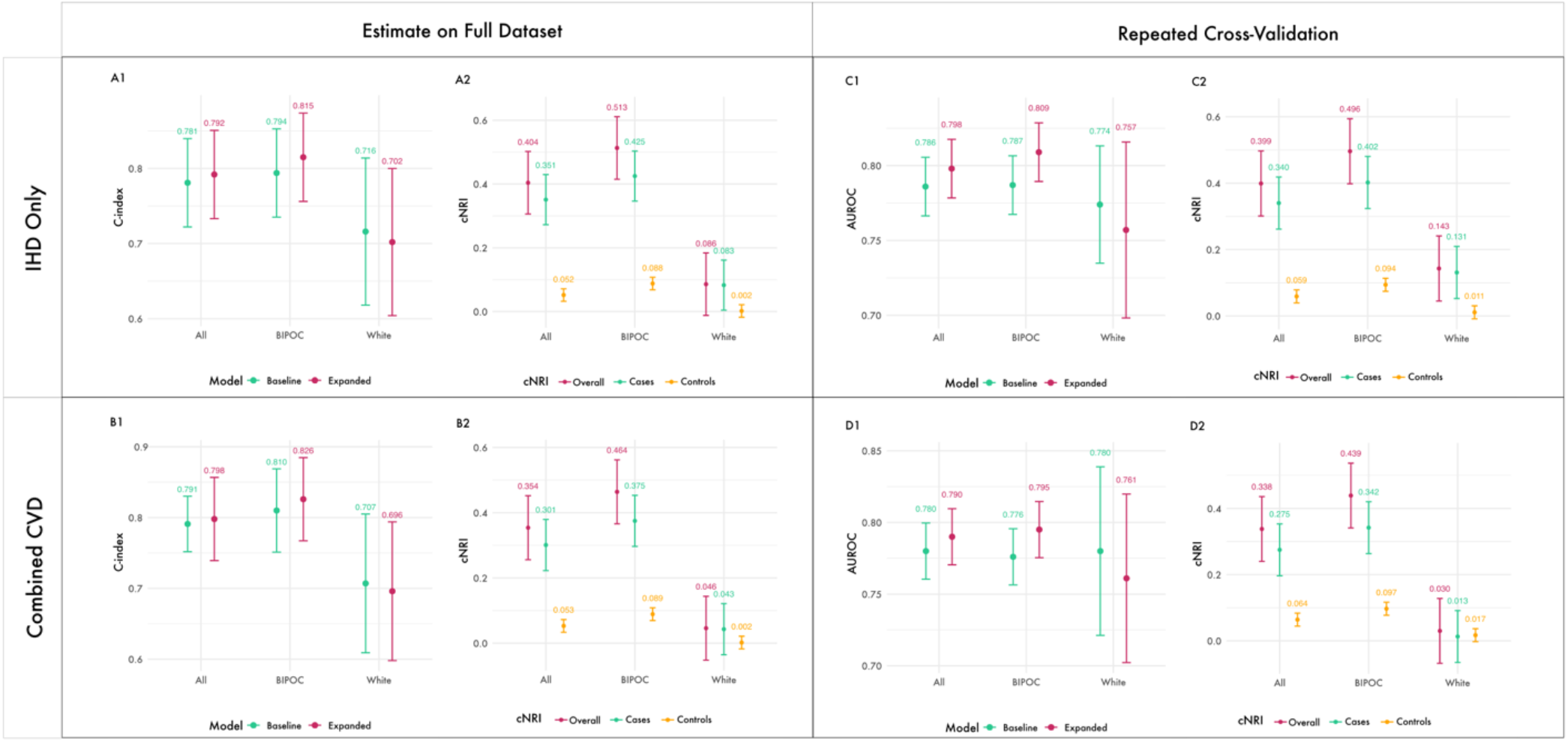
Evaluation of Dysmenorrhea as a Predictor in IHD and CVD Risk Models. Panel A-B: Comparison of baseline and expanded Cox regression models for predicting ischemic heart disease (IHD) (A1-2) and combined CVD risk (B1-2) in the MS LLM-EHR cohort. The baseline model uses the Framingham Offspring 30-year cardiovascular disease risk score (30FRS) as a predictor, while the expanded model includes dysmenorrhea as an additional predictor. Panel C-D: Repeated 5x5 cross-validation results for IHD (C) and combined CVD (D) prediction using area under the receiver operating characteristic curve (AUROC) and cNRI as performance metrics

Adding dysmenorrhea significantly improved the goodness-of-fit of prognostic Cox regression models over 30FRS risk score alone, both when predicting IHD (LRT p-value <0.001) and combined CVD (LRT p-value <0.001). Moreover, the inclusion of dysmenorrhea enhanced the model’s discriminative power (C-index 0.781 vs 0.792 for IHD prediction and 0.791 vs 0.798 for combined CVD prediction). The cNRI was overall positive (0.40 in IHD prediction and 0.35 in combined CVD prediction), primarily driven by improvements in reclassifying cases, with less pronounced benefits for net reclassification in controls. The highest benefit from including dysmenorrhea was observed for BIPOC patients (C-index 0.794 vs 0.815 and cNRI 0.51 for IHD prediction, C-index 0.810 vs 0.826 and cNRI 0.46 for combined CVD prediction).

Additionally, we performed a repeated cross-validation experiment to empirically estimate the discriminative benefit of adding dysmenorrhea to a baseline IHD prediction model using 30FRS as a predictor (Figure 4, Panel C), as well as for combined CVD prediction (Figure 4, Panel D) using area under the receiving operator curve (AUROC) and cNRI as metrics. For both IHD and combined CVD prediction, including dysmenorrhea as a predictor significantly increased the AUROC (0.786 vs 0.798, p- value 0.019 for IHD, 0.780 vs 0.790, p-value 0.002 for combined CVD), especially in BIPOC patients (0.787 vs 0.809, p-value <0.001 for IHD, 0.776 vs 0.795, p-value<0.001 in combined CVD). However, including dysmenorrhea led to invariant performance for White patients (AUROC of 0.774 vs 0.757, p- value=0.169) for IHD prediction and poorer performance for combined CVD prediction (0.780 vs 0.761, p-value=0.014).

## Discussion

In this study we investigated the risk for IHD in women with dysmenorrhea across three diverse cohorts totaling 251,264 patients. We report three main findings. Firstly, we confirmed findings from a previous study^17^ showing increased HR with doubling of the risk of IHD in women with dysmenorrhea independently from other cardiovascular or reproductive risk factors, corroborating them across cohorts. Secondly, we showed that the effect of dysmenorrhea is moderated by race and ethnicity, with increased risk found mainly in BIPOC women, in particular Asian and Hispanic women. Thirdly, we demonstrate that dysmenorrhea is an age-dependent risk factor, particularly important for patients between 30-40 years with persistent dysmenorrhea since adolescence/emerging adulthood. We finally empirically confirm the prognostic value of dysmenorrhea for improved cardiovascular risk stratification by showing increased discrimination power and improved reclassification of patients when adding dysmenorrhea as a predictor to standard cardiovascular risk models, especially when used to estimate risk in BIPOC patients.

To the best of our knowledge, there was only one previous work by Yeh et al.^17^ where the authors investigated the risk of IHD in women with dysmenorrhea using a nation-wide insurance claim dataset, identifying an increased HR for IHD in this population. While their study made an important contribution to the field, it relied solely on coded diagnoses to define dysmenorrhea cases and controls, which can introduce limitations. We expand upon their findings by demonstrating that coded diagnoses alone may not fully capture the complexity of dysmenorrhea, as evidenced by a false negative rate of up to 37.3% when identifying controls. In our study, we confirm their results in the AoU cohort, where dysmenorrhea status was similarly ascertained via coded diagnoses alone. Additionally, we show that these findings also hold when cases and controls are more accurately defined, namely using our phenotyping algorithm in the MS LLM-EHR cohort, as well as defining it via self-reported measures in the ALSWH-S5 and ALSWH cohorts. Moreover, Yeh et al. did not adjust for other reproductive health factors, such as irregular menstrual cycles, heavy menstrual bleeding, and polycystic ovary syndrome, which could potentially influence the association between dysmenorrhea and IHD. In our analysis, we adjust for these variables and demonstrate that dysmenorrhea remains an independent risk factor for IHD. Finally, whereas Yeh et al. excluded patients with secondary causes of dysmenorrhea, we find that both primary and secondary dysmenorrhea are associated with an increased risk of IHD across our cohorts.

Several mechanisms might be involved in linking dysmenorrhea to increase risk of IHD. Firstly, dysmenorrhea is associated with ischemic pain^19^ through the release of prostaglandin and the induced myometrial contractions, as well as altered perfusion^20^. Prostaglandins, in turn, have been linked to increased risk of IHD^21,22^, and the molecular mechanisms responsible for altered perfusion in uterine tissue might parallel similar effects on myocardial tissue. Moreover, dysmenorrhea patients have alterations in autonomic nervous system associated with sympathetic hyperactivation^13,14^, a well-known cardiovascular risk factor^23,24^. Dysmenorrhea is also associated with increased stress^15,16^, which itself is a cardiovascular risk factor, particularly for young women^25^ and women of minoritized racial and ethnic groups^26,27^. Finally, secondary causes of dysmenorrhea, and in particular endometriosis have previously been associated with increased cardiovascular risk^28^. While in our work we show that dysmenorrhea remains a significant risk factor for IHD in the absence of secondary causes, we cannot exclude undiagnosed endometriosis as an underlying effect mechanism, given the well-known delay in diagnosis, with a considerable proportion of endometriosis patients remaining undiagnosed throughout their lifetime^29^. It is important to note, however, that secondary dysmenorrhea typically arises after 25 years of age^30^. Our results, on the other hand, suggest that dysmenorrhea arising before 25 years of age and persisting across the thirties is mostly associated to increased IHD risk compared to early or late dysmenorrhea only, therefore suggesting an involvement of primary dysmenorrhea disease mechanisms that adds risk beyond secondary causes that might develop. In this regards it was recently suggested that disease mechanisms of primary dysmenorrhea might favor the development of endometriosis in later years^31^, possibly complicating further the unequivocal distinction between primary and secondary dysmenorrhea, and contributing to the idea that shared pathophysiological mechanisms might underly the increased cardiovascular risk seen for both primary and secondary dysmenorrhea cases.

Our secondary analyses indicate a heterogeneous effect of dysmenorrhea on IHD risk across race and ethnicity groups. Specifically, we observed a significant interaction between race and ethnicity and dysmenorrhea, with BIPOC patients showing higher HRs compared to White patients. Notably, race and ethnicity-specific sub-analyses suggest that dysmenorrhea may serve as a stronger IHD risk factor in Hispanic and Asian populations compared to others. Sensitivity analysis on AoU (Appendix VI) found no significant confounding by socio-economic variables (e.g., education, income, insurance type), suggesting that the observed interaction between race and dysmenorrhea in predicting IHD risk is independent of these factors. Considering that previous studies have reported a higher prevalence of severe dysmenorrhea in Hispanic and Asian women compared to Black/African American and White patients^32,33^, it is possible that the severity—rather than the mere presence—of dysmenorrhea plays a critical role in increasing IHD risk. This suggests that future research should explore more precise definitions of dysmenorrhea, particularly focusing on severity and subtypes, to better understand its potential link to cardiovascular risk. It is, however, important to note that while this interaction showed a consistent trend across cohorts, it was not statistically significant across all cohorts, and the confidence intervals for group-specific estimates exhibited considerable overlap. Thus, future studies are necessary to replicate and confirm these findings.

Improving cardiovascular risk stratification for young women is particularly important given the recent incidence and mortality rate trends showing a stagnation for this demographic in comparison to their older and male counterpart^34–36^. To this aim, identifying novel female-specific risk marker that can be assessed at a younger age is crucial. In this study we show that dysmenorrhea is not only a risk factor for IHD, but also particularly an important one for younger women between 30 and 40 years of age. These findings are paralleled by similar findings with respect to BIPOC, where dysmenorrhea is a significant risk factor more so for BIPOC rather than White patients. Again, this subpopulation is also showing greater disparities in cardiovascular outcomes^26,37^. Altogether, these results suggest that dysmenorrhea is a particularly interesting risk marker that appears well-suited for improving risk stratification in this subpopulation. Indeed, our exploratory analyses reveal that adding dysmenorrhea as a risk marker improves discrimination, goodness-of-fit and reclassification indices of prognostic models over an established risk equation developed specifically for the younger population. However, this seems to come at the expenses of a slight decrease in performance in White patients, and further research is needed to determine how to best integrate dysmenorrhea in clinical risk assessment.

Although our findings contribute to the understanding of dysmenorrhea and its effect on long-term cardiovascular health, they are subject to some limitations that warrant discussion. First, while we excluded patients with known menopausal status from our cohort, the inability to reliably ascertain peri- menopausal states remains a limitation. Given that the hormonal fluctuations associated with the peri- menopausal transition can significantly influence cardiovascular risk, this factor should ideally be accounted for in future analyses. Additionally, our determination of several key covariates, including the outcome of interest (IHD), relied on coded diagnoses and self-reported data. Although IHD is generally well-documented, there is potential for underreporting or miscoding. To enhance the accuracy and specificity of future research, it is recommended that studies explore the associations of dysmenorrhea using more precise definitions of IHD and its subtypes. Moreover, for secondary analyses we aggregate race and ethnicity groups into BIPOC and White self-reported race. This approach was necessary to achieve sufficient statistical power for our analyses, but we acknowledge the limitations of aggregating distinct racial and ethnic identities into broader categories, and future research should ensure enough power to characterize the heterogeneity of dysmenorrhea’s association to IHD across more granular racial and ethnic categories.

Gynecologists and cardiologists must increasingly collaborate to enhance women’s cardiovascular health^38^, as more reproductive- and female-specific cardiovascular risk factors are being investigated for effective stratification and prevention. Our research contributes to this effort by highlighting the role of dysmenorrhea in identifying patients at increased risk for IHD. However, we also demonstrate that dysmenorrhea is often overlooked, rarely coded as a diagnosis in EHR and infrequently mentioned in clinical notes by physicians. Additionally, dysmenorrhea is seldom included as a variable in large longitudinal studies on women’s health, with the notable exception of the Australian Longitudinal Study on Women’s Health. These findings are not unexpected, given the historical neglect of dysmenorrhea in both research and clinical settings^12^. Our results indicate that treating dysmenorrhea, especially in the absence of secondary causes, as a ’normal’ condition associated with menstruation may overlook significant health mechanisms. We urge researchers and physicians to increase their awareness of dysmenorrhea, ensuring its proper documentation during clinical encounters and inclusion in large-scale longitudinal studies on women’s health. This will ultimately provide the necessary tools to understand the complex relationship between menstrual conditions and cardiovascular health, fostering the essential collaboration between gynecologists and cardiologists and improving patient outcomes.

## Methods

### Overall Study Design

The main objective of this work was to investigate the risk of IHD in women with dysmenorrhea. To this aim, we analyzed and compared the results from three distinct cohorts (See Figure 1), two EHR-based retrospective cohorts, namely the Mount Sinai (MS) EHR and the All of Us cohort, as well as a prospective cohort, namely the Australian Longitudinal Survey on Women’s Health (ALSWH). Importantly, the three cohorts are characterized by differences in how dysmenorrhea status is ascertained, informing us on the reproducibility of the findings across different exposure definitions. For AoU, cases were defined as a history of SNOMED coded diagnosis of dysmenorrhea prior or at enrollment. In the MS cohort, dysmenorrhea status was similarly ascertained via SNOMED-coded diagnoses alone, but the index date for follow-up corresponded to a random diagnosis event for cases (MS Full cohort). A curated sub-cohort (MS LLM-EHR cohort) was additionally built using information stored in clinical notes together with SNOMED-coded diagnoses to identify cases and controls with known dysmenorrhea status. This enabled us to directly compare the use of coded diagnosis alone versus more refined dysmenorrhea status characterization for identifying correlations with future IHD risk. For the ALSWH, dysmenorrhea was ascertained through self-reported measures of menstrual pain across survey timepoints. The whole ALSWH cohort was analyzed with time-varying covariates updated at each available survey, while a separated ALSWH-S5 sub-cohort was created by using the 5^th^ survey as the index date for follow-up, where age range is comparable to the MS EHR, MS LLM-EHR and AoU cohorts.

Secondary aims of the study were to ascertain heterogeneity in dysmenorrhea effects on IHD across racial groups and across temporal and age patterns of dysmenorrhea, as well as the prognostic value of dysmenorrhea over established cardiovascular risk markers.

Each secondary analysis was performed on all cohorts and sub-cohorts that enabled such characterization.

This study was approved by the Icahn School of Medicine at Mount Sinai’s Institutional Review Board (Institutional Review Board 19-00951). The IRB has determined that this research involves no greater than minimal risk and approved the waiver for informed consent.

### Datasets

A full description of the EHR codes and methods used for covariate extraction can be found in Appendix I.

#### Mount Sinai (MS)

Mount Sinai Data Warehouse (MSDW) is a OMOP common data model-mapped dataset of EHR data from the Mount Sinai Health System, the largest hospital system in the New York metropolitan area containing more than 10 million patient records. For the Mount Sinai cohort, we extracted all EHRs of female-born patients with at least one record of a routine gynecological examination between 15 and 45 years of age. We excluded all patients with a history of hysterectomy or premature ovarian failure. According to how dysmenorrhea status was ascertained, we define two different MS sub-cohorts: MS- EHR, where dysmenorrhea status was determined via SNOMED-coded diagnosis alone, and MS-LLM- EHR, where dysmenorrhea status was determined via a large language model (LLM)-driven phenotyping algorithm using additional information stored within clinical free text (see below). As index dates for start of observation we selected a random diagnosis date of dysmenorrhea for cases and a randomly sampled routine gynecological encounter between 15 and 45 years of age for controls. Patients were considered followed up until the last available EHR entry. We extracted baseline covariates including current or previous coded diagnoses of endometriosis, uterine leiomyoma (fibroids), irregular menstrual cycles (IM), heavy menstrual bleeding (HMB), polycystic ovary syndrome (PCOS), migraine, diabetes mellitus (DM), hypertension (HTN), hypertensive disorders of pregnancy (HDP) and gestational diabetes (GDM). Moreover, we extracted information regarding smoking status, use of hormonal contraceptives (HCU) and BMI if present within 1 year from the index date. Finally, we extracted self-reported race/ ethnicity (Black or African American, Asian, Hispanic, White, and Others – including American Indian/Alaskan Natives and Hawaiian Natives/Pacific Islander or Mixed Race). We used a separate category to code patients without self-reported race. Outcome of interest was ischemic heart disease as defined by SNOMED code ‘Ischemic Heart Disease (414545008) and its descendant concepts. A full description of the covariate extraction process can be found in Appendix I.

All patients with a first IHD diagnosis occurring before or at the index date were excluded from the analyses, as well as patients with a history of hysterectomy or premature ovarian insufficiency/early menopause.

#### All of Us (AoU)

The All of Us dataset^39^ consists of a multi-center OMOP-mapped EHR dataset linked to baseline surveys collected at enrollment as well as genetic and wearable sensor data. The date of baseline survey completion was used as index date for start of follow-up.

We included all female-born patients that completed the baseline survey and reported having menstrual periods. We collected covariates and IHD outcome from EHR records with the same rationale used for MS dataset except for history of hypertension, diabetes, endometriosis, fibroids, PCOS and IHD which was defined as either a previous coded diagnosis, or a self-reported history of in the Personal Health History (PHH) Survey.

#### Australian Longitudinal Survey on Women’s Health

The Australian Longitudinal Survey on Women’s Health^40^ (ALSWH) is a longitudinal population-based survey first conducted in 1996 and with a total of 8 survey-related data available across 22 years (S1- S8). We used the data from the 1973-1978 cohort (age range 18-50). The survey includes questions on frequency of severe menstrual pain, irregular menstrual cycles and heavy menstrual bleeding. Response options were ‘never’, ‘rarely’, ‘sometimes’, and ‘often’, which we dichotomized into present when ‘sometimes’ or ‘often’ was reported, and absent otherwise. Moreover, information regarding hormonal contraceptive use, smoking status, new or past diagnoses of endometriosis, migraine, hypertension, diabetes and BMI was used to define the available covariates. Due to inconsistent capture of information regarding PCOS, hypertensive disorder of pregnancy and gestational diabetes mellitus across surveys, we could not unbiasedly include them in the dataset. The outcome of ischemic heart disease was defined as self-reported diagnosis of or treatment for ‘Heart Disease (including heart attack, angina)’. ALSWH dataset contains repeated measurements at different survey timepoint. The full ALSWH cohort was built to include time-varying covariates, where each covariate was sequentially updated across surveys when new values became available, and the outcome status ascertained at the next available survey point. Patients reporting a history of ‘Heart Disease’ prior to S1, as well as those reporting a history of hysterectomy and/or bilateral oophorectomy at S4-5 were excluded from the analyses. For each patient, the follow-up ended at either the first mention of ‘Heart Disease’, or the last available survey.

For improved comparability with the MS and AoU cohorts, we included a sub-cohort ALSWH-S5 where we used the 5^th^ ALSWH survey (S5) data as index point, where the age range (31-37, mean 33) is closest to the mean age at index date for MS-Full (34), MS LLM-EHR (33) and AoU (35). We used information from S1-S5 for assessing covariate status, namely history of hypertension, diabetes, endometriosis and migraine, as well as S5-only to establish current dysmenorrhea, IM, HMB status, smoking, HCU and BMI. Additionally, information on history of gestational diabetes, hypertensive disorder of pregnancy (S2-4) and PCOS (S4-5) was ascertained for this sub-cohort. Time-to-event was determined based on S6-S8 surveys, with right censoring occurring at the last available survey.

### Dysmenorrhea Phenotyping: MS LLM-EHR Cohort

We curated a subset of the MS cohort dataset (LLM-EHR) where dysmenorrhea status was ascertained using additional information present within physicians’ notes. To this end, we adapted a pre-trained clinical language model (PLM), GatorTron^41^, for classifying gynecological clinical notes for positive, negative or unknown dysmenorrhea status via prompt-based learning with keyword-optimized template insertion^42^. Dysmenorrhea status was then defined following the algorithm depicted in Figure 1. Patients with a SNOMED coded diagnosis and/or at least one clinical note classified as dysmenorrhea were defined as cases. Controls were identified if a patient had at least one clinical note classified as no dysmenorrhea by the PLM and did not meet the case criteria. Those where dysmenorrhea status was not possible to ascertain were defined as unknown and excluded from the MS LLM-EHR dataset. To test performance of the PLM and phenotyping algorithm on our dataset, we randomly selected 200 patients (100 with and 100 without IHD) and performed manual annotation of clinical notes to label dysmenorrhea status. The PLM achieved a macro-averaged F1 score of 0.89 (precision=0.90, recall=0.88). The overall phenotyping algorithm achieved a good performance with a macro-averaged F1 of 0.88 (precision=0.91, recall=0.87). Appendix II describes the details of model development and evaluation. Moreover, in Appendix III we provide proof that the phenotyping algorithm performance is independent from outcome status and therefore does not introduce selection or confounding bias into the analyses.

### Statistical Analyses

#### Propensity Score Matching

Propensity score matching is a statistical technique used to estimate the effect of a factor by accounting for covariates that predict receiving the treatment, thereby reducing selection bias in observational studies. They are used to approximate a randomized controlled study in an observational context, theoretically to infer causal effect estimation^43^. Several approaches for propensity score matching exist, including distance-based matching with subset selection, stratification or a combination of the two. In this work we employ generalized full propensity score matching ^44–46^. This technique assigns every case and control unit to one subclass, minimizing the sum of absolute within-subclass distances to ensure balanced groups and effectively combining distance and stratum matching techniques. It has several advantages over other methods, including a reduced sensitivity to the propensity score model specification^47^, as well as the matching of all subjects without the need to drop unmatched units. We used generalized full propensity score matching implemented in the MatchIt library for R ^48^ to estimate the average marginal effect of dysmenorrhea on IHD accounting for confounding by all covariates, i.e. age, BMI, hypertension, diabetes, smoking status (Ever Smoker/Never Smoker/Unknown), HCU, HMB, IM, endometriosis, fibroids, migraine, PCOS, and race. The propensity score was estimated using a logistic regression of dysmenorrhea on all covariates. We forced exact matching on rounded age to the nearest year. After matching, all standardized mean differences for the covariates were below 0.1 and all standardized mean differences for squares and two-way interactions between covariates were below .15, indicating adequate balance. Full matching uses all treated and all control units, and units were discarded only if no exact matching of rounded age was possible. A detailed description of matching specifications and post-matching covariate balance can be found in Appendix II.

#### Time-To-Event Modeling

We estimated the hazard ratio (HR) for IHD via weighted multivariate cox regression with dysmenorrhea as predictor, the estimated propensity scores as weights, and the identified subclasses as clustering variables enabling robust variance estimation^49^. We additionally fit a weighted regression model including all matching covariates as predictors (Supplementary Material II and Appendix V). Rather than modeling time-to-event using calendar time of follow-up, we used age as a time-scale^50^ accounting for left truncation (for adjusting people entering the study at different ages) as well as right censoring. Modeling on the age scale has several advantages. Age is the strongest single predictor of incident IHD and IHD subtypes, requiring correct modeling of functional relationship between age and IHD. Using age as a timescale obviates this need by implicitly including it in the model specifications^51,52^. Moreover, it improves interpretability of the results while controlling for age-dependent confounding. Finally, it is consistent with some established clinical risk prediction models such as the SCORE^53^ and PREVENT^54^. While the main analyses use age as a time scale, sensitivity to time-scale specification demonstrates no significant differences in the findings (See Appendix V).

Recently, propensity score matching methods have been compared with standard modeling with covariate adjustment showing that choosing the former might introduce less precise effect estimates^55^. We therefore performed additional analyses using standard covariate adjustment by including varying combinations of covariates in unweighted cox regression models. We compared baseline model (Model 1) with dysmenorrhea as the sole covariates to two increasingly adjusted models. Model 2 includes additional cardiovascular risk factors namely hypertension, diabetes, BMI, smoking status and race and ethnicity. Model 3 represents the full model including also reproductive variables endometriosis, fibroids, PCOS, migraine, HCU, IM, HMB as well as pregnancy-related hypertension and gestational diabetes.

For AoU, MS-Full, MS LLM-EHR and ALSWH-S5 datasets a time-to-event model was fit with covariate status established at index date. However, the ALSWH dataset contains repeated measurements at different survey timepoint. We therefore fitted a cox regression model with time-dependent covariates, where all available surveys for each patients were fitted and outcome determined at the next available survey^56^. Because full propensity score matching would introduce bias, we only performed standard covariate adjustment for analyses on ALSWH.

Proportionality assumptions check for cox models as well as additional sensitivity analyses for time-of- enrollment as a time scale can be found in Appendix V.

All analyses were performed with R software 4.3.2. We considered statistical significance at a threshold of 0.05 for p values.

### Secondary Analyses

#### Effect Modification by Race and Ethnicity

We assessed effect heterogeneity across racial and ethnic groups on the AoU, MS Full and MS LLM- EHR cohorts, since race and ethnicity information was not available in the ALSWH. We performed our main analyses using a binary variable for aggregate racial and ethnic groups, where White patients were compared to Black, Native and People of Color (BIPOC) patients. BPOC included Asian, Black or African American, Hispanics, Native Hawaiian/Other Pacific Islander, Native American/Alaskan Natives, and Middle Eastern/Norther African. We first performed full propensity score matching with exact matching on binarized race and ethnicity and rounded age to the next year, while including distinct racial and ethnic groups in the propensity score model to ensure fine-grained balance of race and ethnicity across the cohort. Post-matching balance metrics are reported in Appendix IV. We then estimated the moderation effect by fitting a weighted cox regression with an interaction term between dysmenorrhea and dichotomized race and ethnicity. We finally estimated stratum-specific HR across White and BIPOC groups via stratified weighted cox regression across binarize race and ethnicity. We finally estimated fine- grained race and ethnicity-specific HR via weighted stratified cox regression across distinct racial and ethnic groups. We also estimated the interaction term and the strata-specific estimates without matching via adjusted cox regression including all covariates as predictors. Because race and ethnicity are social constructs that might reflect differences in socio-economic variables, we performed additional sensitivity analyses to assess whether the association modification as well as direct association between dysmenorrhea and IHD was confounded by socio-economic variables (See Appendix VI).

#### Effect Modification by Secondary Causes of Dysmenorrhea

Dysmenorrhea is broadly categorized as primary or secondary dysmenorrhea, according to the presence of secondary causes such as endometriosis or fibroids. To assess the influence of secondary causes of dysmenorrhea on the heterogeneity of the effect estimate, we first performed a moderation analysis for presence of secondary causes (endometriosis or fibroids). We first performed full propensity score matching with exact matching on secondary causes, achieving very good balance across secondary causes strata. Post-matching balance metrics are reported in Appendix IV. We subsequently fit a weighted cox regression for the interaction between dysmenorrhea and secondary causes to estimate a significant moderation effect. We then estimated strata-specific HR for dysmenorrhea on IHD via stratified weighted cox regression. We additionally performed analyses for comparable unweighted adjusted cox regression without propensity score matching.

#### Effect Heterogeneity Across Age Groups

To assess if the effect of dysmenorrhea is heterogeneous across age, we first analyzed the ALSWH cohort to estimate the time-varying coefficient β(*t*) for dysmenorrhea across age in a multivariate cox regression model including all covariates and using age as a time scale. First, we visually explored the time-varying coefficient estimated via Cox-Snell residuals analysis using the cox.zph function in the R Survival package^57^. We then fitted a second cox model with a quadratic time transformation of the dysmenorrhea variable ((*f*(*x*, *t*) = *x*(*t* − 35)^2^, where 𝑥 is the dysmenorrhea indicator and *t* is the time in years on age as a scale), which corresponds to the functional form visualized through cox-snell residual analysis. Given the statistically significant coefficient for the time-transformed dysmenorrhea variable, we confirm the inverse-U shaped temporal pattern. We finally estimated β(*t*) as a step function over age groups <=30, >30-<=40 and >40 by first breaking the dataset into time-dependent parts using the SurvSplit function in the R Survival package^57^, followed by estimation of stratum-specific HR via stratified multivariate cox regression (i.e. piecewise regression).

To externally validate these findings, we estimate the stratum-specific HR for age-groups >30-<=40 vs <=30 or >40 within the MS-Full and MS LLM-EHR cohorts. These were estimated via stratified weighted logistic regression following full propensity score matching with exact matching on age at index date rounded to the nearest integer.

#### Natural History Patterns of Dysmenorrhea

Dysmenorrhea is a condition that can vary in intensity across the reproductive lifespan. Different natural history patterns of dysmenorrhea might therefore underly different pathophysiological mechanisms with heterogeneous effects on risk of IHD. To assess a differential effect across natural history patterns of dysmenorrhea we analyzed the ALSWH-S5 cohort. We defined three natural history patterns of dysmenorrhea: persistent dysmenorrhea, which arises before 25 years of age and persists at index date (i.e. 31-37 years); early dysmenorrhea, where dysmenorrhea arises before 25 but does not persist at index date; and late dysmenorrhea, where dysmenorrhea was absent before 25 years of age and is present at index date. We excluded all patients for which dysmenorrhea status before 25 years could not be ascertained due to missingness. We dummy-coded this multiclass definition of dysmenorrhea pattern and estimated the HR over no dysmenorrhea ever via weighted cox regression following full propensity score matching as well as in a multivariate cox regression model adjusted for all available covariates. We additionally estimated the HR for persistent dysmenorrhea compared to other natural history patterns via multivariate cox regression adjusted for all available covariates by restricting the analyses on patients with a history of dysmenorrhea only (N= 3361).

Additionally, we estimated the effect of age at first diagnosis in the AoU cohort. Because the AoU cohort’s index date corresponds to the date of baseline survey completion, the cohort’s cases represent patients with a history of dysmenorrhea. We estimate the effect of age at diagnosis for cases only by fitting a cox regression with age at diagnosis as a predictor, and adjusted for history of endometriosis and fibroids given the relationship between secondary causes and age at first diagnosis^58^. We then estimated the HR for age at first diagnosis before and after 25 years of age compared to controls by dummy-coding a 3- class variable (no dysmenorrhea, dysmenorrhea before 25, dysmenorrhea after 25) via multivariate cox regression adjusting for all available covariates.

#### Prognostic Value of Dysmenorrhea

To determine the added prognostic value of dysmenorrhea over established cardiovascular risk factors we firstly compared the addition of dysmenorrhea as a predictor to a cox regression model including the risk index calculated from the 30-year CVD Framingham Risk Score^59^ (30FRS). The 30FRS is a 30-year risk calculator developed on the Framingham Offspring cohort^59^ aged 20-59 for CVD outcomes including IHD, stroke, heart failure, atrial fibrillation and peripheral vascular disease. We choose this risk model since it was shown to correlate better with CVD risk in younger patients compared to other established risk models such as the pooled cohort equation^36^.

For this experiment we used the MS LLM-EHR cohort. We extracted additional features systolic blood pressure (SBP) and prescription of antihypertensive treatment from the EHR, which, together with age, BMI, smoking and diabetes were required as inputs to the risk model. Moreover, we extracted additional outcomes needed to estimate the overall cardiovascular event incidence, namely stroke, heart failure, atrial fibrillation and peripheral vascular disease using SNOMED coded diagnoses. A detailed description of covariate extraction is described in Appendix II.

We compared the discriminative performance of both baseline (30FRS Risk index alone) and expanded (30FRS risk index + dysmenorrhea) models via concordance index (C-index) and continuous net reclassification improvement (cNRI). To compare the goodness-of-fit between models, we performed log-likelihood ratio tests. We further estimated C-index and cNRI for White and BIPOC patients, separately.

To empirically validate the predictive value of dysmenorrhea for IHD prediction we conducted a repeated cross-validation experiment to compare a baseline IHD prediction model using the 30FRS calculated risk alone vs an expanded model additionally including dysmenorrhea. The models where trained to predict IHD event at censoring time. We performed a 5-fold cross-validation repeated five times, where for each split two separated logistic regression models were trained on the training data and evaluated on the left-out fold. The baseline logistic regression model included the IHD risk at censoring time calculated from the 30FRS, age, time-since-index-date, the interaction between age and time-since-index-date, and the quadratic transformation of time-since-index-date as covariates. The expanded model included additional terms for dysmenorrhea and the interaction between dysmenorrhea and time-since-index-date. The IHD risk at censoring time was defined as the cumulative risk was estimated using the risk equations and baseline risk estimates provided by the authors of the 30FRS study^59^. We additionally performed the same experiment for combined CVD. We compared the baseline and expanded models using the area under the receiver operating characteristic curve (AUROC) and cNRI. We estimated the average performance metrics and 95% confidence intervals across cross-validation folds. To evaluate whether the observed differences in AUROC between the baseline and expanded models were statistically significant, we employed a paired two-sided Wilcoxon signed-rank test. Metrics were additionally estimated for BIPOC and White patients, separately.

## Data Availability

The Australian Longitudinal Survey on Women’s Health Core Dataset is available on the Australian Data Archive (https://ada.edu.au/australian-longitudinal-study-on-womens-health-alswh/) upon approval.

The All of Us registered tier dataset is available upon approval at https://workbench.researchallofus.org. The Mount Sinai EHR dataset is available upon request and after Mount Sinai approval.

## Code Availability

Code for building cohorts and reproducing the analyses presented in this manuscript is accessible at github.com/eugenial/Dysmenorrhea_IHD

## Data Availability

The Australian Longitudinal Survey on Women's Health Core Dataset is available on the Australian Data Archive (https://ada.edu.au/australian-longitudinal-study-on-womens-health-alswh/) upon approval.
The All of Us registered tier dataset is available upon approval at https://workbench.researchallofus.org.
The Mount Sinai EHR dataset is available upon request and after Mount Sinai approval.

## Acknowledgments

We thank the Scientific Computing and Data group at the Icahn School of Medicine at Mount Sinai for the computational and data resources, and the IT staff expertise supported by the Clinical and Translational Science Awards (CTSA) grant UL1TR004419 from the National Center for Advancing Translational Sciences. The research on which this paper is based was conducted in part as part of the Australian Longitudinal Study on Women’s Health by the University of Queensland and the University of Newcastle. We are grateful to the Australian Government Department of Health and Aged Care for funding and to the women who provided the survey data. We gratefully acknowledge *All of Us* participants for their contributions, without whom this research would not have been possible. We also thank the National Institutes of Health’s *All of Us* Research Program for making available the participant data and cohort examined in this study. This work was supported by the Hasso Plattner Foundation and the AIR.MS project.

## Abbreviations

30FRS: 30-Year Framingham Risk Score

ALSWH: Australian Longitudinal Study on Women’s Health AoU: All of Us

ASCVD: Atherosclerotic Cardiovascular Disease AUROC: Area Under the Receiver Operating Curve BIPOC: Black, Indigenous and Other People of Color C-Index: Concordance Index

cNRI: Continuous Net Reclassification Index DM: Diabetes Mellitus

EHR: Electronic Health Record GDM: Gestational Diabetes Mellitus

HDP: Hypertensive Disorders of Pregnancy HMB: Heavy Menstrual Bleeding

HR: Hazard Ratio HTN: Hypertension

IHD: Ischemic Heart Disease IM: Irregular Menstruation LRT: Log-Likelihood Ratio Test MS: Mount Sinai

## References

1. Vos, T. et al. Global burden of 369 diseases and injuries in 204 countries and territories, 1990–2019: a systematic analysis for the Global Burden of Disease Study 2019. The Lancet 396, 1204–1222 (2020).

2. Roth, G. A. et al. Global Burden of Cardiovascular Diseases and Risk Factors, 1990-2019: Update From the GBD 2019 Study. J. Am. Coll. Cardiol. 76, 2982–3021 (2020).

3. Martin, S. S. et al. 2024 Heart Disease and Stroke Statistics: A Report of US and Global Data From the American Heart Association. Circulation 149, e347–e913 (2024).

4. Vervoort, D. et al. Addressing the Global Burden of Cardiovascular Disease in Women: JACC State-of-the-Art Review. J. Am. Coll. Cardiol. 83, 2690–2707 (2024).

5. Henry, S., Bond, R., Rosen, S. E., Grines, C. & Mieres, J. H. Challenges in Cardiovascular Risk Prediction and Stratification in Women. Cardiovasc. Innov. Appl. 3, 329 (2019).

6. Jørstad, H. T. et al. Estimated 10-year cardiovascular mortality seriously underestimates overall cardiovascular risk. Heart 102, 63–68 (2016).

7. Arnett, D. K. et al. 2019 ACC/AHA Guideline on the Primary Prevention of Cardiovascular Disease: A Report of the American College of Cardiology/American Heart Association Task Force on Clinical Practice Guidelines. Circulation 140, e596–e646 (2019).

8. Adedinsewo, D. A. et al. Cardiovascular Disease Screening in Women: Leveraging Artificial Intelligence and Digital Tools. Circ. Res. 130, 673–690 (2022).

9. Lo, A. C. Q., Lo, C. C. W. & Oliver-Williams, C. Cardiovascular disease risk in women with hyperandrogenism, oligomenorrhea/menstrual irregularity or polycystic ovaries (components of polycystic ovary syndrome): a systematic review and meta-analysis. Eur. Heart J. Open 3, oead061 (2023).

10. Dubey, P., Reddy, S., Singh, V., Yousif, A. & Dwivedi, A. K. Association of heavy menstrual bleeding with cardiovascular disease in US female hospitalizations. BMC Med. 22, 208 (2024).

11. Keenan, K., Hipwell, A. E. & Polonsky, T. S. Menstrual Cycle Irregularity in Adolescence Is Associated With Cardiometabolic Health in Early Adulthood. J. Am. Heart Assoc. 12, e029372 (2023).

12. Iacovides, S., Avidon, I. & Baker, F. C. What we know about primary dysmenorrhea today: a critical review. Hum. Reprod. Update 21, 762–778 (2015).

13. Oladosu, F. A. et al. Persistent autonomic dysfunction and bladder sensitivity in primary dysmenorrhea. Sci. Rep. 9, 2194 (2019).

14. Wang, Y.-J., Wang, Y.-Z. & Yeh, M.-L. A Prospective Comparison Study of Heart Rate Variability During Menses in Young Women With Dysmenorrhea. Biol. Res. Nurs. 18, 465–472 (2016).

15. Yamamoto, K., Okazaki, A., Sakamoto, Y. & Funatsu, M. The relationship between premenstrual symptoms, menstrual pain, irregular menstrual cycles, and psychosocial stress among Japanese college students. J. Physiol. Anthropol. 28, 129–136 (2009).

16. Wang, L. et al. Stress and dysmenorrhoea: a population based prospective study. Occup. Environ. Med. 61, 1021–1026 (2004).

17. Yeh, C.-H., Muo, C.-H., Sung, F.-C. & Yen, P.-S. Risk of Ischemic Heart Disease Associated with Primary Dysmenorrhea: A Population-Based Retrospective Cohort Study. J. Pers. Med. 12, 1610 (2022).

18. Yeh, C.-H., Sung, F.-C., Muo, C.-H., Yen, P.-S. & Hsu, C. Y. Stroke Risk in Young Women with Primary Dysmenorrhea: A Propensity-Score-Matched Retrospective Cohort Study. J. Pers. Med. 13, 114 (2023).

19. Sen, E., Ozdemir, O., Ozdemir, S. & Atalay, C. R. The Relationship between Serum Ischemia- Modified Albumin Levels and Uterine Artery Doppler Parameters in Patients with Primary Dysmenorrhea. Rev. Bras. Ginecol. E Obstet. Rev. Fed. Bras. Soc. Ginecol. E Obstet. 42, 630–633 (2020).

20. Dmitrović, R. Transvaginal color Doppler study of uterine blood flow in primary dysmenorrhea. Acta Obstet. Gynecol. Scand. 79, 1112–1116 (2000).

21. Beccacece, L., Abondio, P., Bini, C., Pelotti, S. & Luiselli, D. The Link between Prostanoids and Cardiovascular Diseases. Int. J. Mol. Sci. 24, 4193 (2023).

22. Hirsh, P. D., Campbell, W. B., Willerson, J. T. & Hillis, L. D. Prostaglandins and ischemic heart disease. Am. J. Med. 71, 1009–1026 (1981).

23. Curtis, B. M. & O’Keefe, J. H. Autonomic Tone as a Cardiovascular Risk Factor: The Dangers of Chronic Fight or Flight. Mayo Clin. Proc. 77, 45–54 (2002).

24. Boudou, N. et al. Direct evidence of sympathetic hyperactivity in patients with vasospastic angina. Am. J. Cardiovasc. Dis. 7, 83–88 (2017).

25. Vaccarino, V. et al. Sex Differences in Mental Stress-Induced Myocardial Ischemia in Patients With Coronary Heart Disease. J. Am. Heart Assoc. 5, e003630.

26. Kalinowski, J., Taylor, J. Y. & Spruill, T. M. Why Are Young Black Women at High Risk for Cardiovascular Disease? Circulation 139, 1003–1004 (2019).

27. Felix, A. S. et al. Stress, Resilience, and Cardiovascular Disease Risk Among Black Women. Circ. Cardiovasc. Qual. Outcomes 12, e005284 (2019).

28. Couto, C. P. do, Policiano, C., Pinto, F. J., Brito, D. & Caldeira, D. Endometriosis and cardiovascular disease: A systematic review and meta-analysis. Maturitas 171, 45–52 (2023).

29. Husby, G. K., Haugen, R. S. & Moen, M. H. Diagnostic delay in women with pain and endometriosis. Acta Obstet. Gynecol. Scand. 82, 649–653 (2003).

30. Proctor, M. & Farquhar, C. Diagnosis and management of dysmenorrhoea. BMJ 332, 1134–1138 (2006).

31. Clemenza, S. et al. Is primary dysmenorrhea a precursor of future endometriosis development? Gynecol. Endocrinol. Off. J. Int. Soc. Gynecol. Endocrinol. 37, 287–293 (2021).

32. Banikarim, C., Chacko, M. R. & Kelder, S. H. Prevalence and Impact of Dysmenorrhea on Hispanic Female Adolescents. Arch. Pediatr. Adolesc. Med. 154, 1226–1229 (2000).

33. Deuster, P. A., Powell-Dunford, N., Crago, M. S. & Cuda, A. S. Menstrual and Oral Contraceptive Use Patterns Among Deployed Military Women by Race and Ethnicity. Women Health 51, 41–54 (2011).

34. Minissian, M. B. et al. Ischemic Heart Disease in Young Women. J. Am. Coll. Cardiol. 80, 1014–1022 (2022).

35. Wilmot, K. A., O’Flaherty, M., Capewell, S., Ford, E. S. & Vaccarino, V. Coronary Heart Disease Mortality Declines in the United States From 1979 Through 2011: Evidence for Stagnation in Young Adults, Especially Women. Circulation 132, 997–1002 (2015).

36. An, J. et al. Incidence of Atherosclerotic Cardiovascular Disease in Young Adults at Low Short-Term But High Long-Term Risk. J. Am. Coll. Cardiol. 81, 623–632 (2023).

37. Smilowitz, N. R., Maduro, G. A., Lobach, I. V., Chen, Y. & Reynolds, H. R. Adverse Trends in Ischemic Heart Disease Mortality among Young New Yorkers, Particularly Young Black Women. PLoS ONE 11, e0149015 (2016).

38. Brown, H. L. et al. Promoting Risk Identification and Reduction of Cardiovascular Disease in Women Through Collaboration With Obstetricians and Gynecologists: A Presidential Advisory From the American Heart Association and the American College of Obstetricians and Gynecologists. Circulation 137, e843–e852 (2018).

39. null, null. The “All of Us” Research Program. N. Engl. J. Med. 381, 668–676 (2019).

40. Dobson, A. J. et al. Cohort Profile Update: Australian Longitudinal Study on Women’s Health. Int. J. Epidemiol. 44, 1547–1547f (2015).

41. Yang, X. et al. A large language model for electronic health records. Npj Digit. Med. 5, 1–9 (2022).

42. Alleva, E., et al. Keyword-optimized Template Insertion for Clinical Information Extraction via Prompt- based Learning. Preprint at 10.48550/arXiv.2310.20089 (2023).

43. Austin, P. C. An Introduction to Propensity Score Methods for Reducing the Effects of Confounding in Observational Studies. Multivar. Behav. Res. 46, 399–424 (2011).

44. Hansen, B. B. Full Matching in an Observational Study of Coaching for the SAT. J. Am. Stat. Assoc. (2004) doi:10.1198/016214504000000647.

45. Rubin, D. B. & Thomas, N. Combining Propensity Score Matching with Additional Adjustments for Prognostic Covariates. J. Am. Stat. Assoc. (2000).

46. Stuart, E. A. & Green, K. M. Using Full Matching to Estimate Causal Effects in Nonexperimental Studies: Examining the Relationship Between Adolescent Marijuana Use and Adult Outcomes. Dev. Psychol. 44, 395 (2008).

47. Austin, P. C. & Stuart, E. A. The performance of inverse probability of treatment weighting and full matching on the propensity score in the presence of model misspecification when estimating the effect of treatment on survival outcomes. Stat. Methods Med. Res. 26, 1654–1670 (2017).

48. Ho, D., et al. MatchIt: Nonparametric Preprocessing for Parametric Causal Inference. (2023).

49. Austin, P. C. The use of propensity score methods with survival or time-to-event outcomes: reporting measures of effect similar to those used in randomized experiments. Stat. Med. 33, 1242–1258 (2014).

50. Korn, E. L., Graubard, B. I. & Midthune, D. Time-to-event analysis of longitudinal follow-up of a survey: choice of the time-scale. Am. J. Epidemiol. 145, 72–80 (1997).

51. Cologne, J. et al. Proportional Hazards Regression in Epidemiologic Follow-up Studies: An Intuitive Consideration of Primary Time Scale. Epidemiology 23, 565 (2012).

52. Thiébaut, A. C. M. & Bénichou, J. Choice of time-scale in Cox’s model analysis of epidemiologic cohort data: a simulation study. Stat. Med. 23, 3803–3820 (2004).

53. SCORE2 working group and ESC Cardiovascular risk collaboration. SCORE2 risk prediction algorithms: new models to estimate 10-year risk of cardiovascular disease in Europe. Eur. Heart J. 42, 2439–2454 (2021).

54. Khan, S. S. et al. Development and Validation of the American Heart Association’s PREVENT Equations. Circulation 149, 430–449 (2024).

55. Elze, M. C. et al. Comparison of Propensity Score Methods and Covariate Adjustment: Evaluation in 4 Cardiovascular Studies. J. Am. Coll. Cardiol. 69, 345–357 (2017).

56. Therneau, T., Crowson, C. & Atkinson, E. Using Time Dependent Covariates and Time Dependent Coffcients in the Cox Model. *cran.r-project.org* https://cran.r-project.org/web/packages/survival/vignettes/timedep.pdf.

57. Therneau, T. M., until 2009), T. L. (original S.->R port and R. maintainer, Elizabeth, A. & Cynthia, C. survival: Survival Analysis. (2023).

58. Osayande, A. S. & Mehulic, S. Diagnosis and Initial Management of Dysmenorrhea.

59. Pencina, M. J., D’Agostino, R. B., Larson, M. G., Massaro, J. M. & Vasan, R. S. Predicting the 30- year risk of cardiovascular disease: the framingham heart study. Circulation 119, 3078–3084 (2009).

