## Appendix for "Risk of Ischemic Heart Disease in Women with Dysmenorrhea: A Longitudinal Analysis of 251,264 Patients Across Three Diverse Cohorts"

### Supplementary Material

#### Appendix I

##### Mount Sinai Cohorts

The Mount Sinai Data Warehouse is an electronic health record (EHR) database mapping EPIC hospital software-derived EHR data into a common data format (OMOP).

We extracted all electronic health records (EHR) from the Mount Sinai Data Warehouse associated with female assigned sex at birth who had at least one encounter for a routine gynecological visit before 45 years of age. Gynecological visits were identified from EPIC-derived source code names listed in Supplementary Material I. All patients with a history of hysterectomy and ovarian failure were excluded from the cohort.

The index date was defined as either a random diagnosis of dysmenorrhea or a random gynecologic encounter for controls. All patients with ischemic heart disease (IHD) diagnoses prior to the index date were excluded from the analyses. Patients were considered followed-up until either the first diagnosis of IHD or the last EHR observation (i.e. last record of either an encounter or a coded diagnosis).

For the Mount Sinai Full Cohort, dysmenorrhea was defined using coded diagnoses alone. We manually revised all EPIC-derived source code names mapped to SNOMED code 'dysmenorrhea' (194696) and all descendant concepts to identify most precise codes. The same procedure was performed for defining history of irregular menstruation and heavy menstrual bleeding, manually selecting source concept codes that were mapped to SNOMED codes 'excessive and frequent menstruation' (197607), 'excessive and frequent menstruation with irregular cycles' (37311979), 'irregular periods' (196168), and 'abnormal menstrual cycle' (4171394). Mapping from source concept names to dysmenorrhea, irregular menstruation and heavy menstrual bleeding definitions are listed in supplementary material I.

For the MS LLM-EHR cohort, dysmenorrhea was defined using the phenotyping algorithm combining coded diagnoses and information stored within clinical notes (see Appendix II, Figure 1).

The IHD outcome was defined as a SNOMED-coded diagnosis of IHD (4185932) and all ancestor concepts excluding old myocardial infarction, myocardial infarction of the newborn and subsequent myocardial infarction. History of endometriosis, fibroids, migraine, PCOS, Hypertension, hypertensive disorders of pregnancy, diabetes mellitus and gestational diabetes mellitus were identified using SNOMED codes listed in the supplementary material I. Smoking status was ascertained using EHR-derived entries stored in the observation table and observed within 1 year from the index date. Hormonal contraceptive use was defined as either a self-reported use of hormonal contraceptive or a recorded
